# Students’ and adult constituents’ perceptions of gender-based violence and feasible prevention programs in high schools in Ho Chi Minh City: A qualitative study

**DOI:** 10.1101/2025.06.23.25330034

**Authors:** Hoa H. Nguyen, Kien G. To, Van TH. Hoang, Lu Gram, Kathryn M. Yount

## Abstract

Gender-based violence (GBV) is a global social problem with adverse health consequences and heightened risk among young people. GBV among high school students is understudied in Vietnam. This study team interviewed 36 students and 64 adult participants to explore perceptions of GBV and the feasibility of prevention programs in three high schools. Physical, psychological, and sexual violence were identified as major forms of GBV. Perceived perpetrator characteristics included “bad blood, impulsive personality,” and perceived survivor characteristics as “quiet, weak” or “talented, good-looking.” “Bad family environments” and gender norms were perceived to contribute to GBV among students. Participants recommended existing school violence prevention programs and developing new GBV interventions that utilize musical performances, film contests, role-playing, or educational games, in collaboration with communities, schools, and families.

## Introduction

Gender-based violence (GBV) is any harm or threat against a person due to gender’s unequal power in society (Lange & Young, 2019; UNICEF, 2025). The risk of GBV is heightened in adolescents, with an estimated lifetime prevalence of 53% for any GBV against girls in school settings across Africa (Beyene et al., 2019). Globally, the 12-month prevalence of violence against children aged 15 to 17 years ranged from 30% in Europe to 58% in North America (Hillis et al., 2016). Exposure to GBV in adolescence and young adulthood is associated with a range of negative health consequences, from death to physical injury and disability, to unwanted pregnancy, and various mental health conditions (Grose et al., 2021; Grose et al., 2019; Nagashima-Hayashi et al., 2022). The harmful consequences of GBV among students may also include poor academic performance (Mingude & Dejene, 2021).

In Vietnam, the estimated prevalence rates of specific forms of GBV against young women are high (Ministry of Labour - Invalids and Social Affairs, 2019). One third of women aged 22 to 78 years faces a high burden of GBV impacting all aspects of their lives (UN Women, 2017), and the prevalence rate of any violent discipline among adolescents (10-14 years) in the month before the Vietnam Sustainable Development Goal Indicators on Children and Women survey was 69% (UNICEF, 2021). A study of sexual misconduct among adolescents 15-19 years in three high schools in Ho Chi Minh City identified high rates of sexual harassment (40.2%), stalking (18.3%), dating violence (13.1%), and sexual violence (8.7%), including higher rates of unwanted sexual attention by staff among boys than girls (18.7% versus 10.9%) and higher rates of dating violence among girls than boys (14.9% versus 9.9%) (Tran et al., 2025). In addition to the personal costs of GBV to survivors, GBV in Vietnam is estimated to cost 1.8% of the country’s Gross Domestic Product annually (UNFPA Vietnam, 2020).

GBV in Vietnam is thought to be rooted in patriarchal norms or gender inequalities in the family and among young people (Bergenfeld, Tamler, et al., 2022; Lewis et al., 2022). Many laws and regulations have been issued to prevent GBV in this normative environment. Illustrative laws include Directive No.18/CT-TTg on strengthening measures to prevent and combat violence against children and child abuse (The Government of Vietnam, 2017b) and Decree No. 80/2017/ND-CP to deter and stop school violence (The Government of Vietnam, 2017a). Despite legal efforts to encourage prevention and response, most female victims (90.4%) do not seek any help from formal service providers (Ministry of Labour - Invalids and Social Affairs, 2019).

Efficacious programs to prevent specific forms of GBV are emerging at the university level in Vietnam (Kathryn M Yount et al., 2023; Lu Gram et al., 2023; Yount et al., 2022); however, programs to prevent GBV in high school students are rare. In a systematic review of GBV prevention studies across Southeast Asia, only seven school-based GBV prevention intervention studies were identified, and the quality of the included intervention studies was poor (Nguyen et al., 2025). Of the seven included intervention studies, only one was a randomized controlled trial, and evaluations mostly have assessed the effectiveness of improving knowledge, attitudes, or skills in school-based programs aiming to prevent sexual violence (Badriah et al., 2023; Nurdin et al., 2018; Santre & Pumpaibool, 2022; Wulanyani et al., 2020). Efficacious programs to prevent GBV in high schools in Vietnam are needed (Kathryn M Yount et al., 2023; Nguyen et al., 2025; Yount et al., 2023).

Understanding the perceptions of students and adult constituents is needed to identify feasible and acceptable programs to prevent GBV in high schools in Vietnam. In the present study, we conducted qualitative research among students and adult constituents in three high schools in Ho Chi Minh City. Three research questions framed the study and analysis:

1. What types of GBV do students and adult constituents (parents, teachers, school managers) perceive to exist among high school students?
2. What characteristics inside and outside the high school environment do participants identify as reasons for GBV?
3. What school-based GBV prevention programs do participants perceive as feasible, acceptable, and effective for high school students?

## Materials and methods

The qualitative study design involved in-depth interviews with high school students (n=36) and focus group discussions with teachers (n=24), parents (n=23), and managers (n=17) affiliated with high schools. The study team followed published guidance regarding the standards for Reporting Qualitative Research for our research checklist (O’Brien et al., 2014).

### Study setting

The study sites were three high schools in Ho Chi Minh City that had different school rankings (low, medium, high) based on the national test scores in 2024 (Dan Tri Newspaper, 2024) and that had participated in a prior quantitative survey of sexual misconduct (Tran et al., 2025). Following this national ranking, the THTH high school was in the top 5 (high level), the NCT high school was ranked 38 (medium level), and the DH high school was ranked 70 (low level) (Dan Tri Newspaper, 2024). All three high schools had classes 10 to 12.

### Sample and recruitment

Eligible participants voluntarily agreed to participate in the study and have the ability to complete the interviews. For in-depth interviews with students, we recruited an equal number of male and female students from classes 10 to 12 at each school (12 students per school and 36 total students). We also invited adult constituents, including parents, school managers, and teachers, for separate focus group discussions at each school (three group discussions per school and nine total focus group discussions).

We used convenience sampling methods to identify and recruit potential participants. Specifically, we met with the Rector of each high school to introduce the study objectives, discuss recruiting participants, and determine the total dates for all interviews and group discussions. The school coordinators provided a list of potential candidates who qualified under our criteria. They then sent invitation letters to prospective participants, arranged interview times, and updated any changes if needed. People who only partially completed the interviews were excluded due to insufficient data for qualitative research. **Table 1** summarizes details of the sample of completed interviews with students and focus group discussions with adult constituents.

**Table 1:** Sample for in-depth interviews and focus group discussions

| **THTH** | **N (F:M)** | **NCT** | **N (F:M)** | **DH** | **N (F:M)** |
| --- | --- | --- | --- | --- | --- |
| ***Students*** |  | ***Students*** |  | ***Students*** |  |
| Class 12 | 4 (2: 2) | Class 12 | 4 (2: 2) | Class 12 | 4 (2: 2) |
| Class 11 | 4 (2: 2) | Class 11 | 4 (2: 2) | Class 11 | 4 (2: 2) |
| Class 10 | 4 (2: 2) | Class 10 | 4 (2: 2) | Class 10 | 4 (2: 2) |
| **Total** | **12 (6:6)** |  | **12 (6:6)** |  | **12 (6:6)** |
| ***Adult constituents*** |  | ***Adult constituents*** |  | ***Adult constituents*** |  |
| Managers | 1 group (N=5) | Managers | 1 group (N=5) | Managers | 1 group (N=7) |
| Teachers | 1 group (N=8) | Teachers | 1 group (N=8) | Teachers | 1 group (N=8) |
| Parents | 1 group (N=8) | Parents | 1 group (N=7) | Parents | 1 group (N=8) |

In-depth interview: Students, Focus group discussion: Adult constituents

**Table 2:**
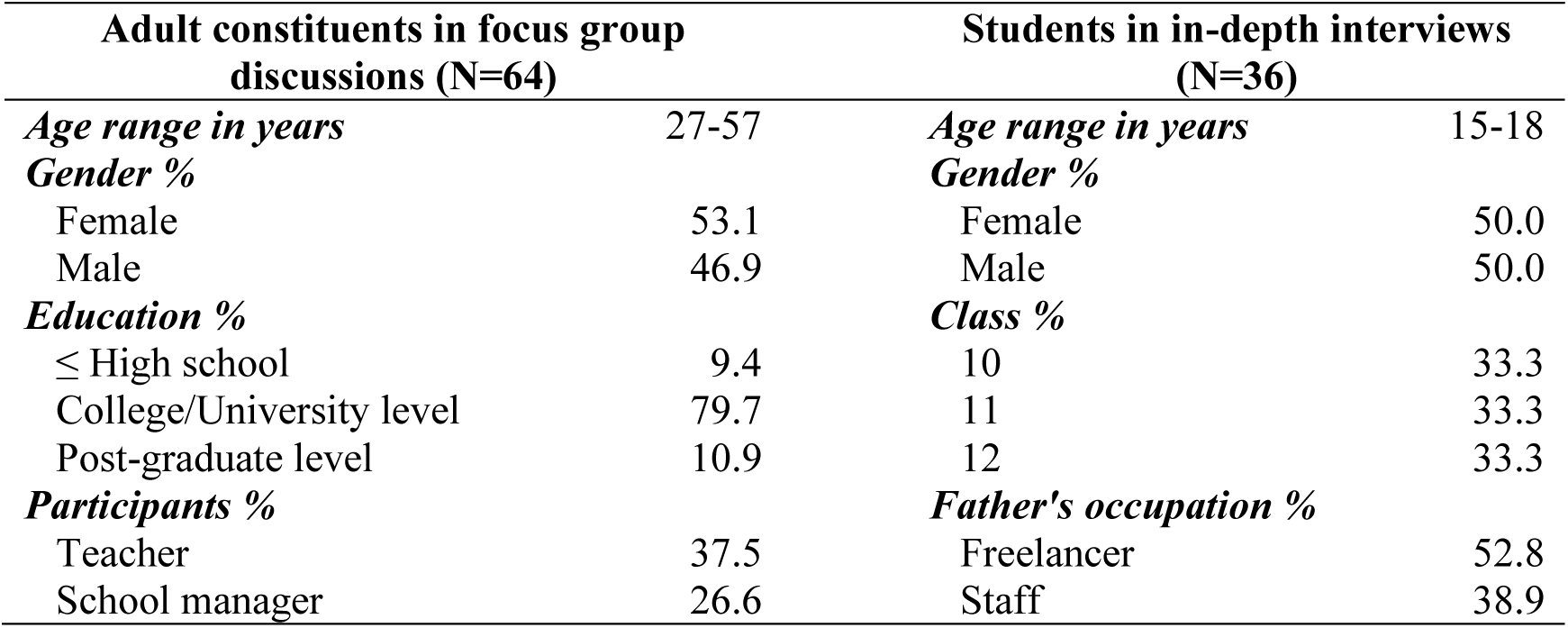
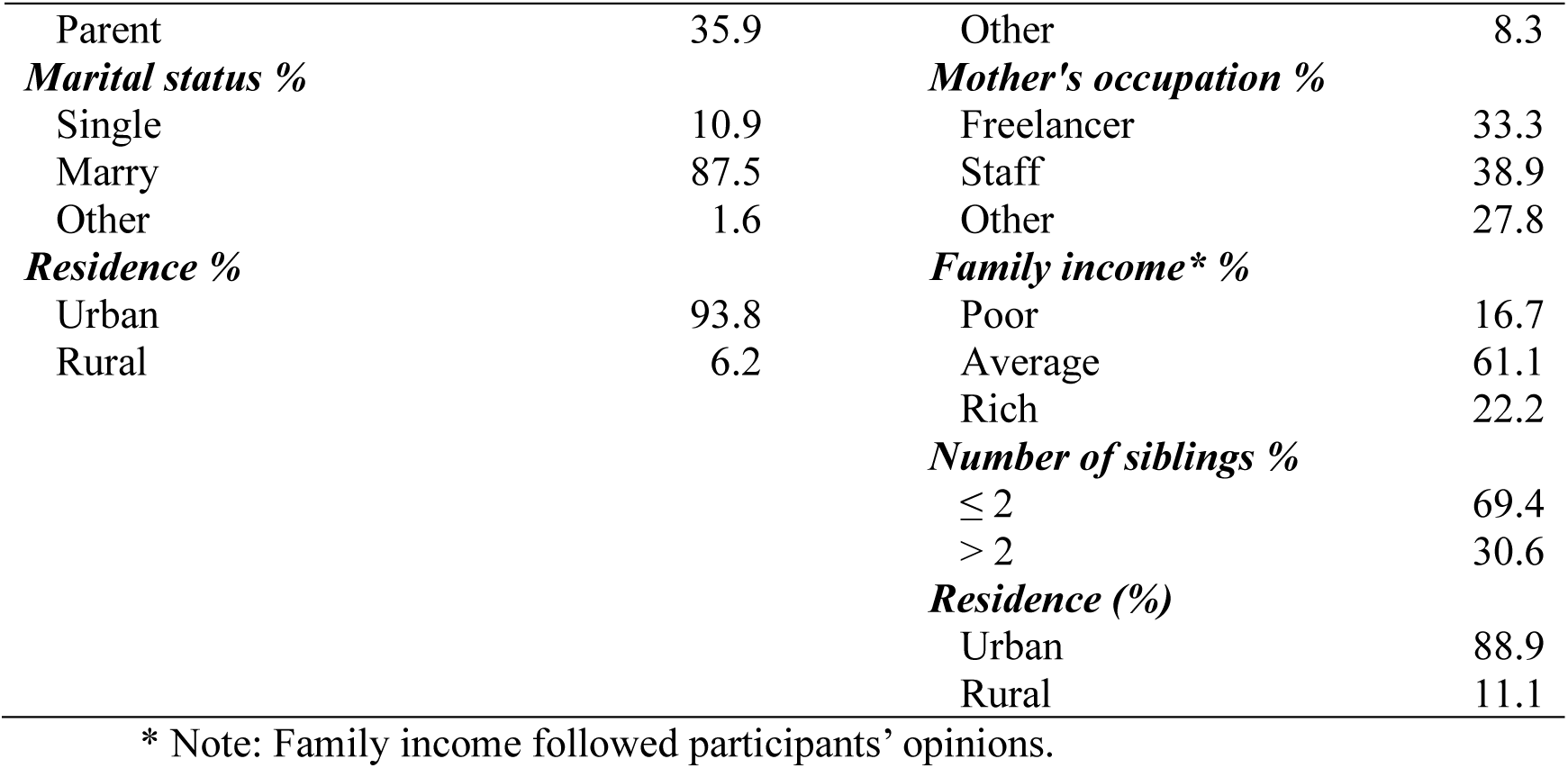
Characteristics of adult constituents and students, three high schools in Ho Chi Minh City

| **Adult constituents in focus group discussions (N=64)** |  | **Students in in-depth interviews (N=36)** |  |
| --- | --- | --- | --- |
| ***Age range in years*** | 27-57 | ***Age range in years*** | 15-18 |
| ***Gender %*** |  | ***Gender %*** |  |
| Female | 53.1 | Female | 50.0 |
| Male | 46.9 | Male | 50.0 |
| ***Education %*** |  | ***Class %*** |  |
| ≤ High school | 9.4 | 10 | 33.3 |
| College/University level | 79.7 | 11 | 33.3 |
| Post-graduate level | 10.9 | 12 | 33.3 |
| ***Participants %*** |  | ***Father's occupation %*** |  |
| Teacher | 37.5 | Freelancer | 52.8 |
| School manager | 26.6 | Staff | 38.9 |
| Parent | 35.9 | Other | 8.3 |
| ***Marital status %*** |  | ***Mother's occupation %*** |  |
| Single | 10.9 | Freelancer | 33.3 |
| Marry | 87.5 | Staff | 38.9 |
| Other | 1.6 | Other | 27.8 |
| ***Residence %*** |  | ***Family income\* %*** |  |
| Urban | 93.8 | Poor | 16.7 |
| Rural | 6.2 | Average | 61.1 |
|  |  | Rich | 22.2 |
|  |  | ***Number of siblings %*** |  |
|  |  | ≤ 2 | 69.4 |
|  |  | > 2 | 30.6 |
|  |  | ***Residence (%)*** |  |
|  |  | Urban | 88.9 |
|  |  | Rural | 11.1 |

\* Note: Family income followed participants' opinions.

### Data collection procedure

#### Interview and group discussion guides

The semi-structured interview guides for students and focus group discussion guides with adult constituents were designed and piloted with students and adults, respectively. We updated the guides based on participants’ feedback. The final student interview guide included five sections: a) a brief introduction to our research, b) warming-up questions with students’ characteristics, c) students’ perspectives on GBV and related factors, and d) future GBV prevention programs for high school students. The final focus group discussion guide for adult constituents included five parts: a) an introduction to our research, b) warming-up questions involving participants’ characteristics, c) perspectives on GBV and related factors, d) current policies to prevent GBV at schools, and e) strategies for future GBV prevention. Overlapping topics across guides allowed us to compare students’ and adults’ group narratives.

#### Fieldwork and data management

Before each interview and group discussion, field staff checked the eligibility of students and adult participants, using the list of candidates provided by school coordinators. The principal investigator and her team conducted all in-depth interviews and focus group discussions in Vietnamese, in person, and in private rooms at the sample schools. Participants could stop the interviews or excuse themselves from the group discussion at any time for any reason. Participants could also skip any questions they did not wish to answer. All interviews and group discussions were recorded using a digital recorder. On average, the interviews lasted 34 minutes (range: 19 - 63 minutes), and the group discussions lasted 66 minutes (range: 41 - 93 minutes). In total, 36 in-depth interviews and nine focus group discussions were conducted from February to March 2024.

Three research assistants transcribed all digital records into Word documents and removed all identifiers. The same research assistants self-checked their transcriptions against the recordings to ensure all information was captured accurately before sending the transcripts to the principal investigator. To ensure accuracy, the principal investigator checked a random one-third sample of the transcriptions against the recordings to ensure completeness and accuracy. The principal investigator saved all recordings and transcriptions on a password-protected drive so that only the research team could access these files.

### Data analysis

All analyses were conducted using the Vietnamese transcripts. The principal investigator imported all transcripts into NVivo software version 14 for coding and data analysis (QSR International, 2023). The principal investigator reviewed the transcripts and drafted a codebook. The codebook included deductive codes derived directly from the interview and discussion guides, as well as inductive codes that emerged during the review and memoing of the transcripts (Fonteyn et al., 2008). The research team reviewed the codebook at regular team meetings and guided revisions to codes and their definitions to ensure completeness and clarity. After that, the principal investigator built coding reports by extracting text segments associated with each code and then synthesized the data using thematic analysis (Kiger & Varpio, 2020) and adapting the Consolidated Framework for Implementation Research for GBV prevention (Yount et al., 2023). Quantitative data on the sample is presented in tabular format, and quotes from the narratives illustrate salient themes that emerged from the thematic analysis.

### Ethical statement

The Board of Ethics in Biomedical Research of the University of Medicine and Pharmacy at Ho Chi Minh City approved the study (No 1244/UMP-BOARD dated 14 December 2023). The principal investigator collected students’ assent and consent forms from adult constituents (parents, teachers, and school managers). The principal investigator obtained verbal informed consent before initiating each in-depth interview and focus group discussion.

## Results

### Characteristics of the sample

A total of 36 students participated in in-depth interviews. An equal number of female and male students participated. Their ages ranged from 15 to 18. The sample also included an equal number of students from classes 10 to 12. Over half of the student participants had fathers working as freelancers, and more than one-third of students had mothers working in staff positions. Almost two-thirds of the student sample (61%) lived in families with an “average” income, and most students (88%) lived in urban areas.

The focus group discussions comprised 64 adults (teachers, school managers, and parents). The age ranges of discussion group participants ranged from 27 to 57. Over half of the adults were female, and nearly 80% held a college or university degree. Over one-third of adult participants were parents (36%) or teachers (38%), and a minority were school managers. Most adults (88%) were married and lived in urban areas (94%).

### Overarching conceptual framework

We adapted the Consolidated Framework for Implementation Research for GBV prevention (Yount et al., 2023), based on narrative evidence from student and adult participants, organizing our qualitative findings into four deductive and inductive thematic domains that emerged from the analysis: a) Perceptions of GBV, b) “Inside” high schools, c) Parental/Family environment, d) “Outside” high school community and society. The views of student and adult participants are compared throughout the results.

**Perceptions about GBV** (Figure 1, Box a)

**Figure 1:**
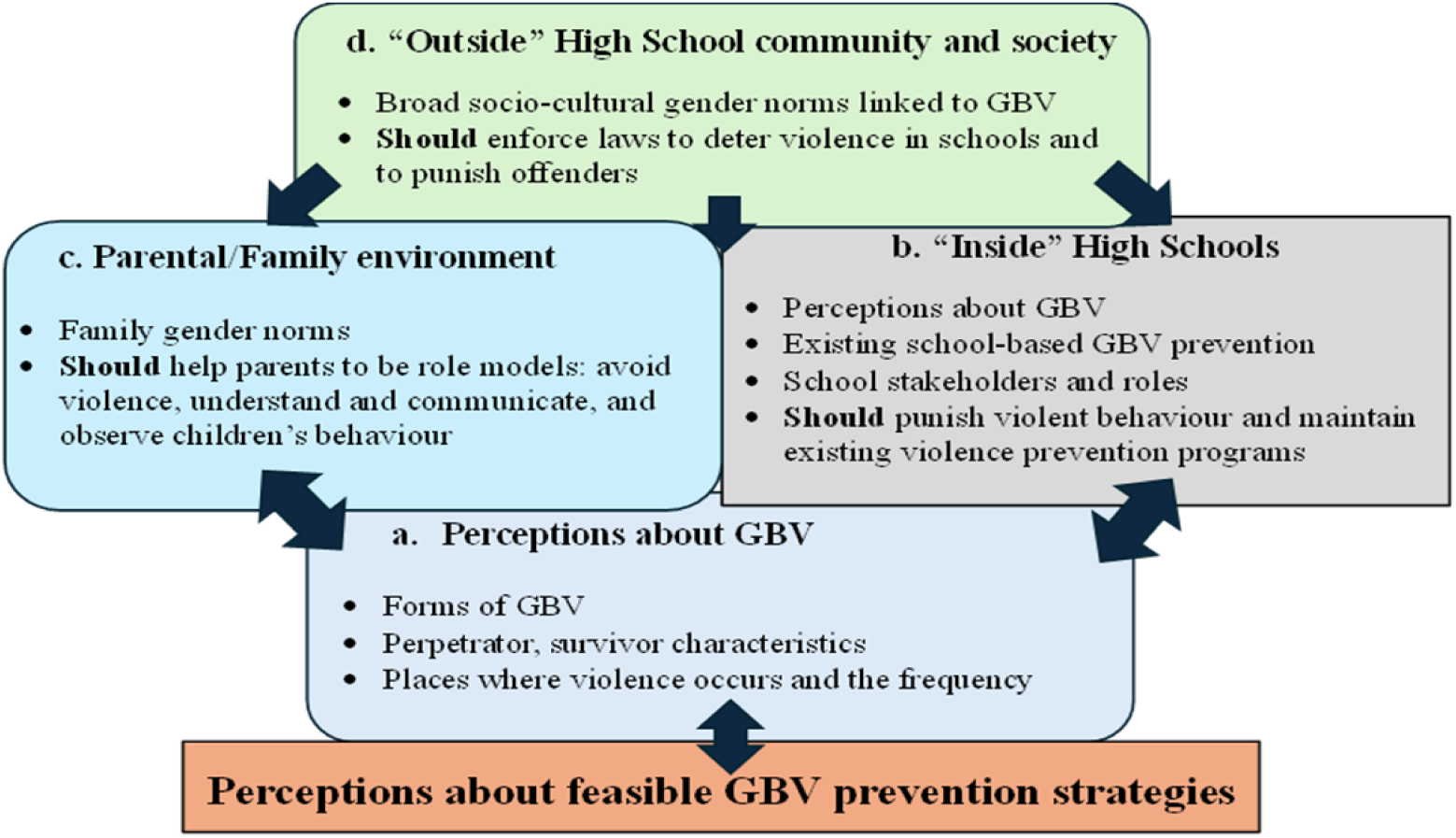
The Consolidated Framework for Implementation Research for GBV prevention in high schools in Ho Chi Minh City

***“Violence is an action”: Perceived forms of GBV in high schools***

Most students and adult participants thought that “violence is an action,” meaning one person enacts different forms of violence against another person. The most common perceived forms of GBV among students and adult participants were physical and psychological violence. Perceived acts of physical violence included attacking, punching, kicking, hitting, or pulling hair, either by one person or a group of people. Perceived acts of psychological violence mostly included using words “to impact others’ psychology” or acts to isolate individuals from others:

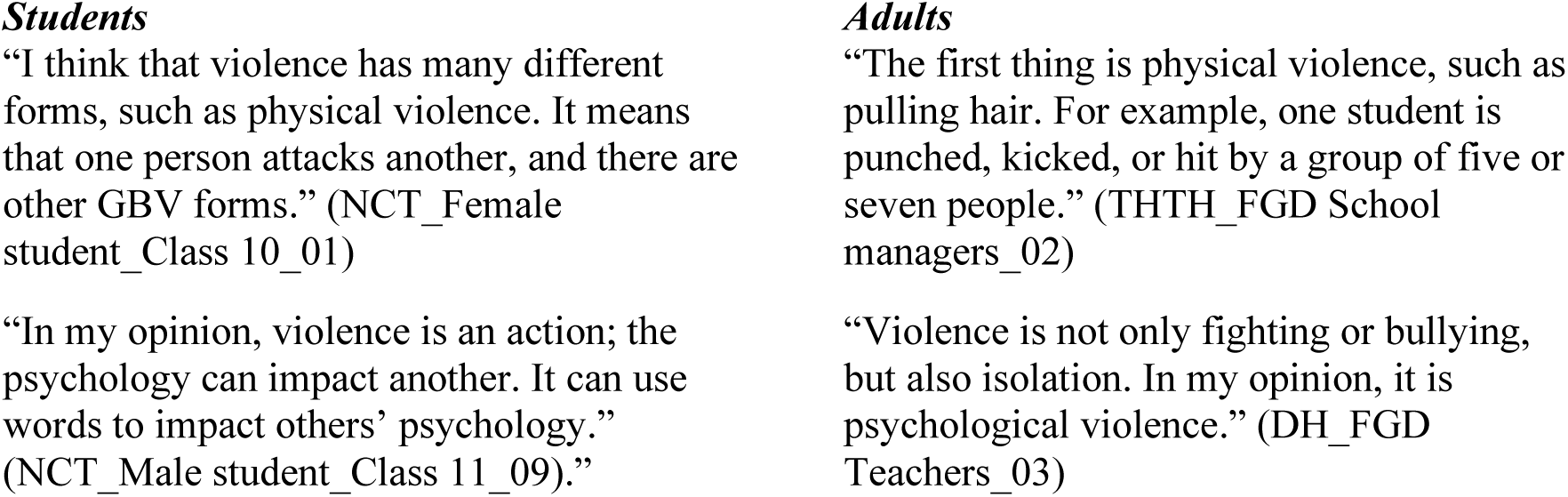

Students and adult participants also mentioned sexual and other forms of violence. Participants often described sexual violence as touching the body or sexual harassment. Other perceived forms of GBV included “being forced to give money” (economic violence) or “doing bad things” more generally:

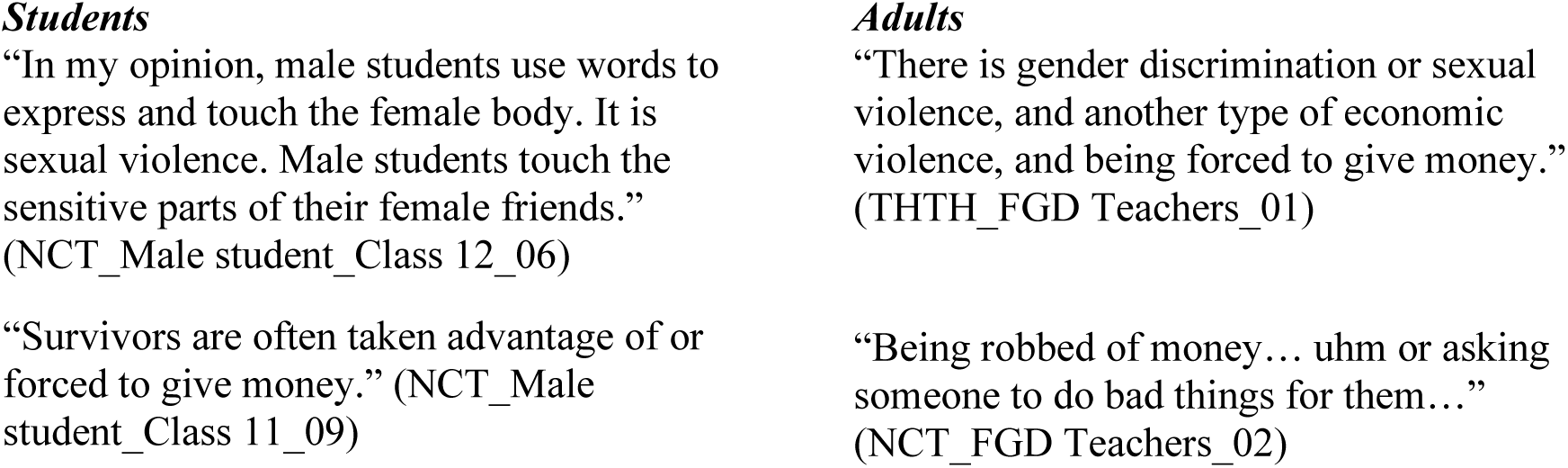

***“Bad blood” and “impulsive personality:” Perceived attributes of students who enact violence***

According to some female students and adult participants, “Bad students” with “bad personalities or tempers” or “bad guys” with “violent blood” tend to be more violent than other students. According to some male students and adult participants, those who commit violence may have an “impulsive personality” or may “show off” or “brag…to show their power” and may “hurt…friends” who refuse to be obedient:

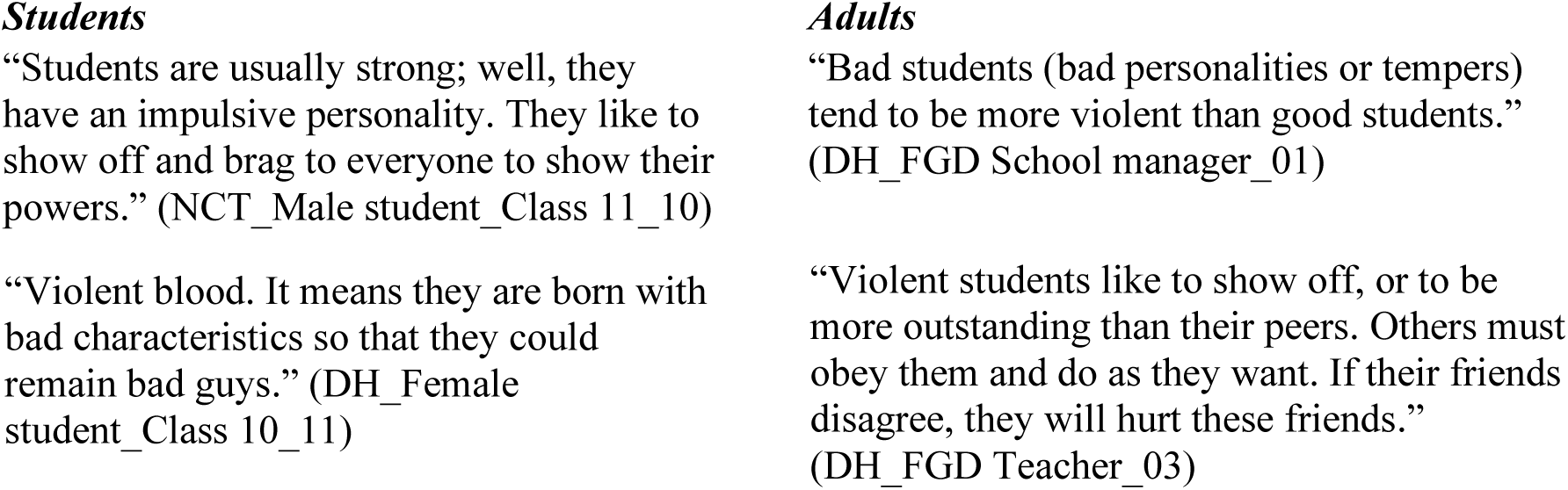

Violent women were described as “petty” or “noisy,” turning small issues into big ones. They also wanted to show off or dominate others. Some people who behaved violently were thought to come from rich families and tried to show their “nobility” by acting violently towards those who were “poorer.”

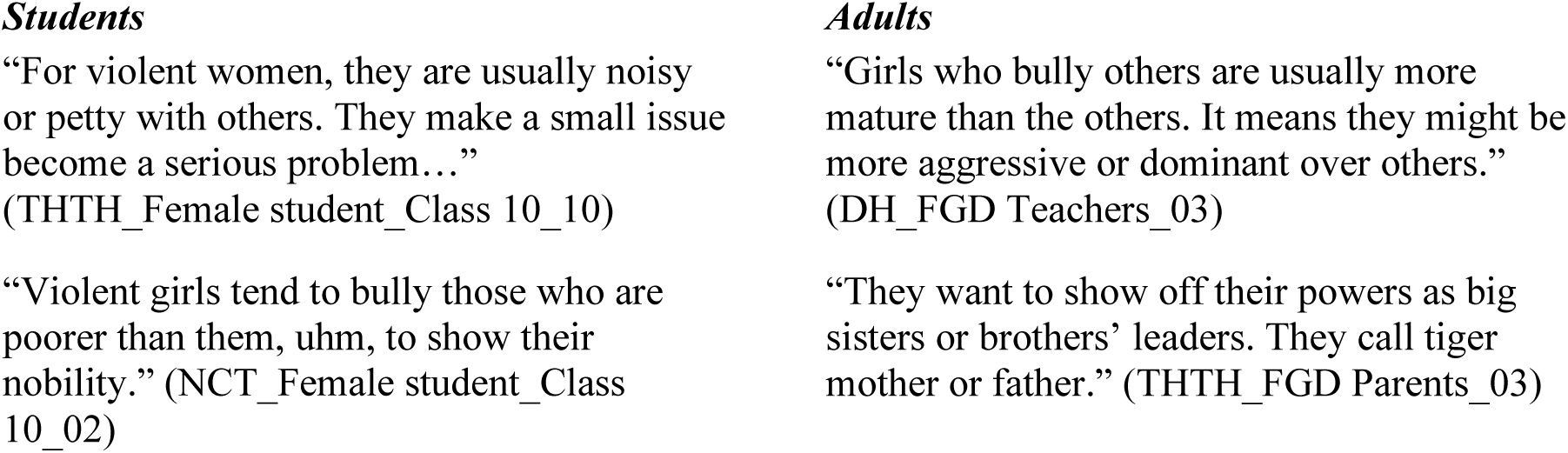

***“Quiet, isolated, shy” or “strong personalities, good performance”: Perceived characteristics of violence survivors***

In general, female or male GBV survivors were not more likely than the other gender to be described as “weak, shy, quiet, bad, or stupid.” However, the attributes of survivors were thought to explain the violence they experienced. For example, having a strong personality was perceived to possibly result in greater social isolation and greater vulnerability to violence:

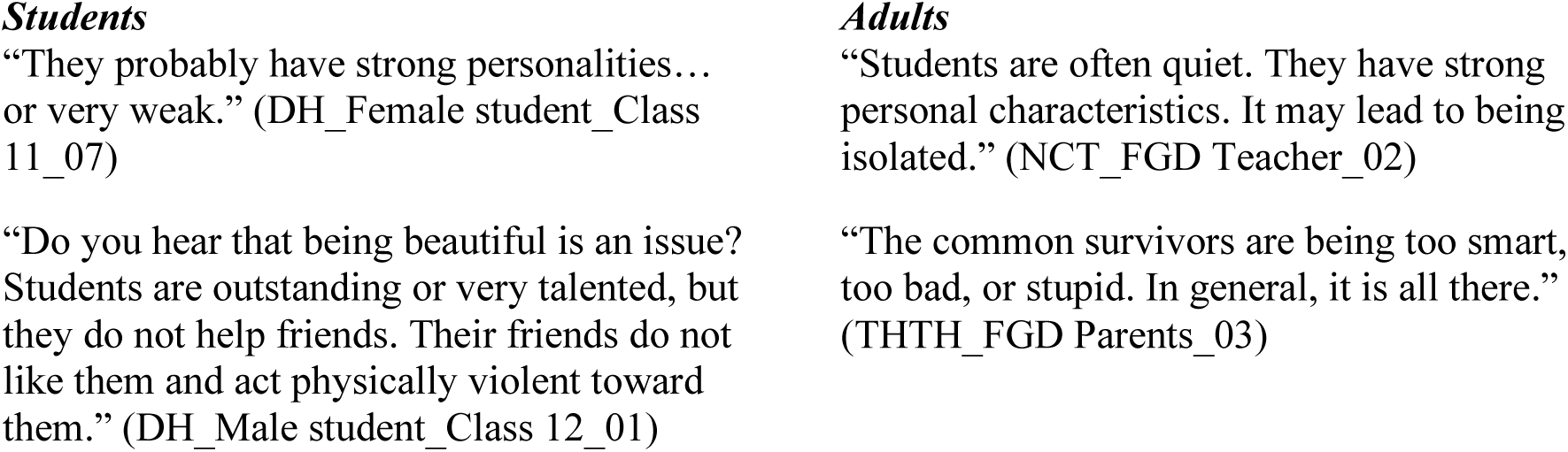

***Perceived places where violence occurs and frequencies***

According to many students and adult participants, violence was perceived to occur in many places without cameras, including in the school gate, as well as in classrooms, toilets, abandoned land, and soccer fields:

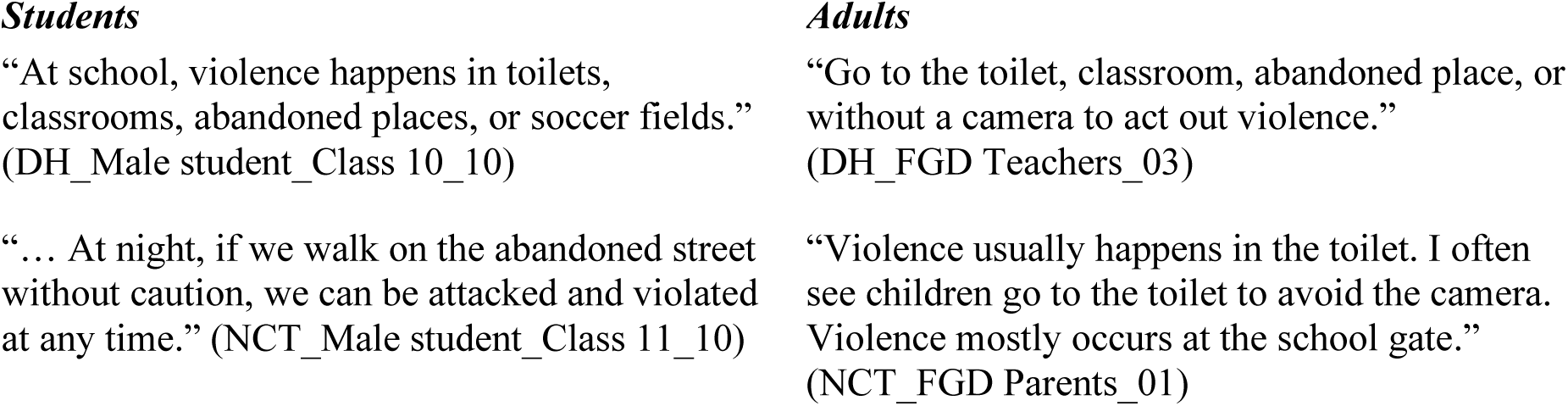

Views differed, however, among student and adult participants about the frequency of GBV. Some students and adults thought that violence occurred every day, whereas other students and adults believed that violence at school was rare:

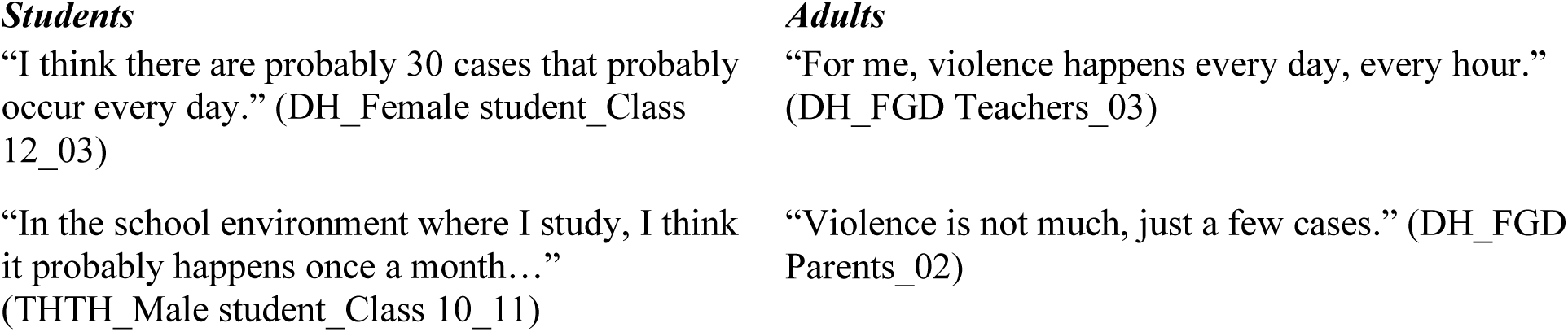

**“Inside” the high school environment influences the occurrence of GBV (**Figure 1**, Box b)**

***There is/is not “gender prejudice” in schools***

Many participants declared no gender discrimination in exam organizations or promoting leadership positions, arguing that male- and female-identified students were treated equally at school. Other participants reported that leadership positions were given to male students:

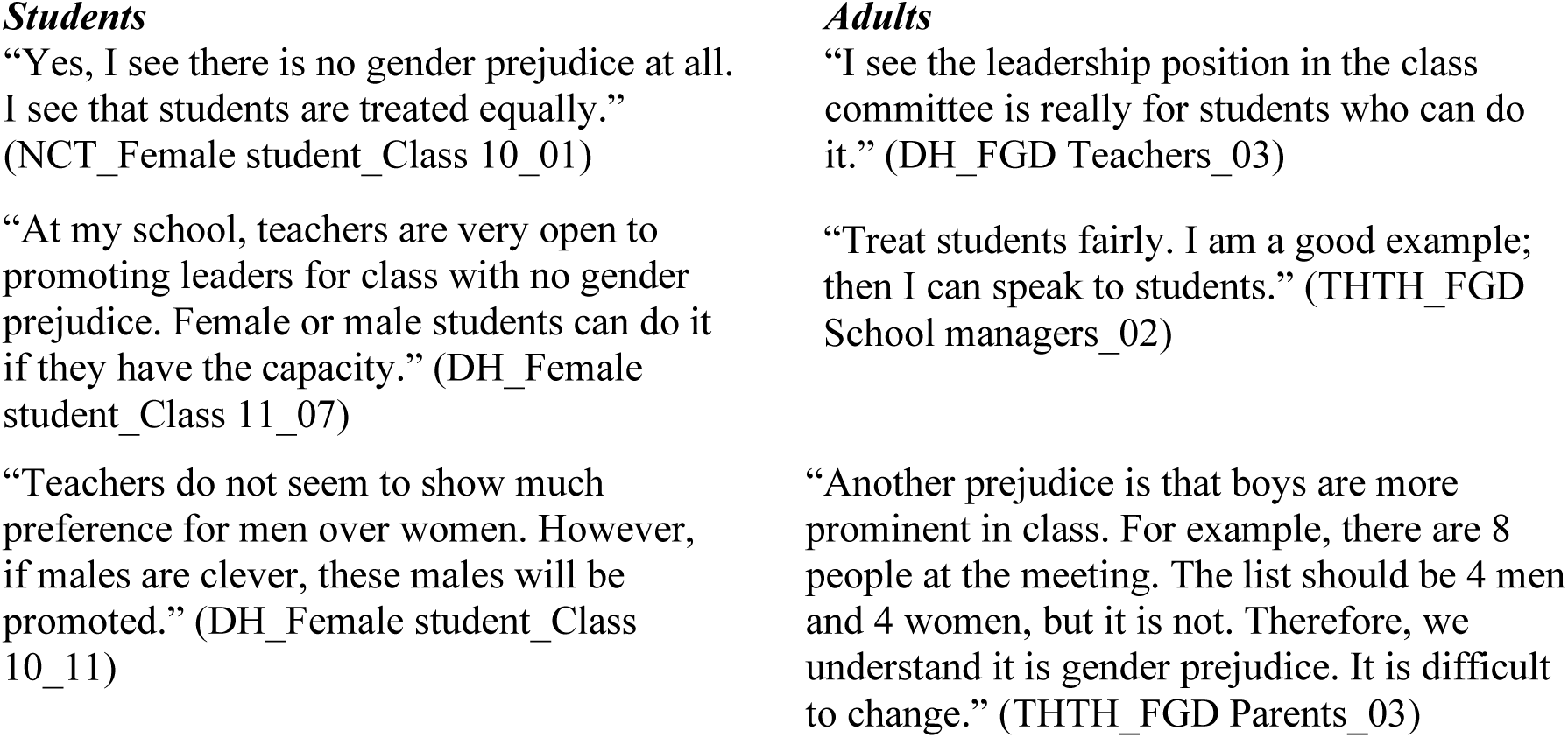

***Available school-based GBV prevention activities***

Most students and adult participants reported that existing school-based prevention programs for high school students provided education on GBV through specific topics at schools. For example, life skills were used to teach the necessary knowledge and skills to prevent violence. GBV prevention was also taught at several weekly Monday activities at all high schools. These programs were not specifically designed to address gender prejudice in schools. They were just a single lesson or a topic taught in the curriculum.

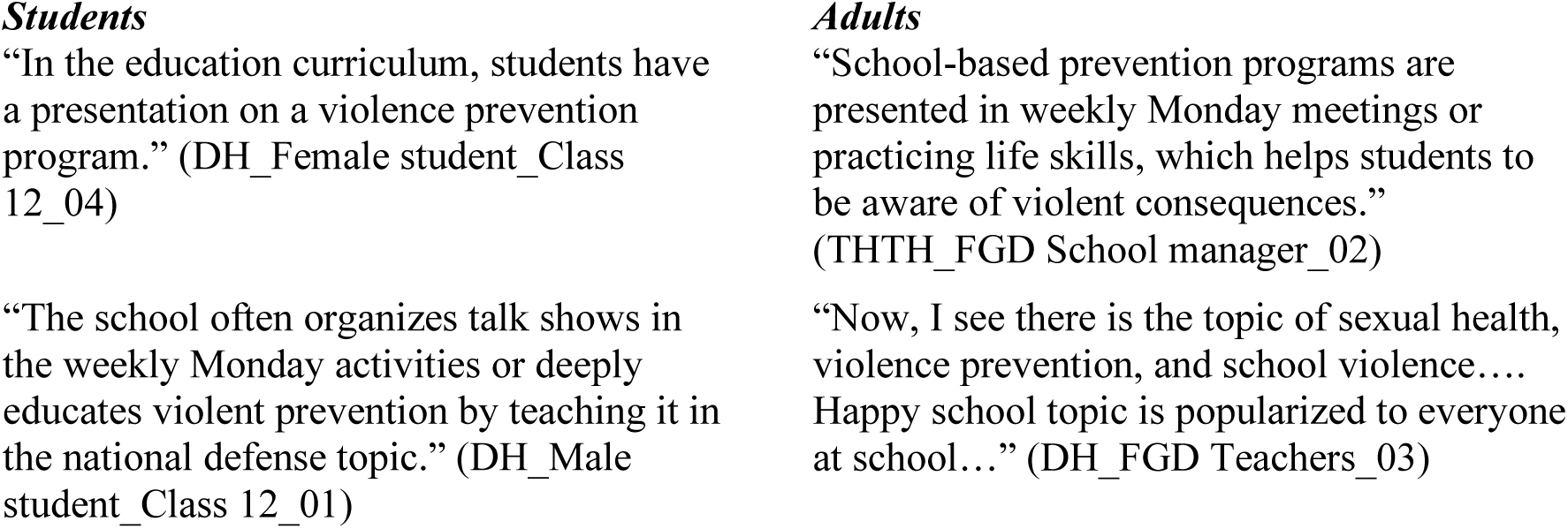

**Family factors relating to GBV** (Figure 1, Box c)

Many students and adult participants stated that family norms contribute to GBV, particularly norms focusing on respecting men more than women and the roles of husbands and wives. According to participants, a husband was customarily thought of as ‘the Lord’ due to his role as the family breadwinner, so the wife was expected to stay at home to do housework or care for children:

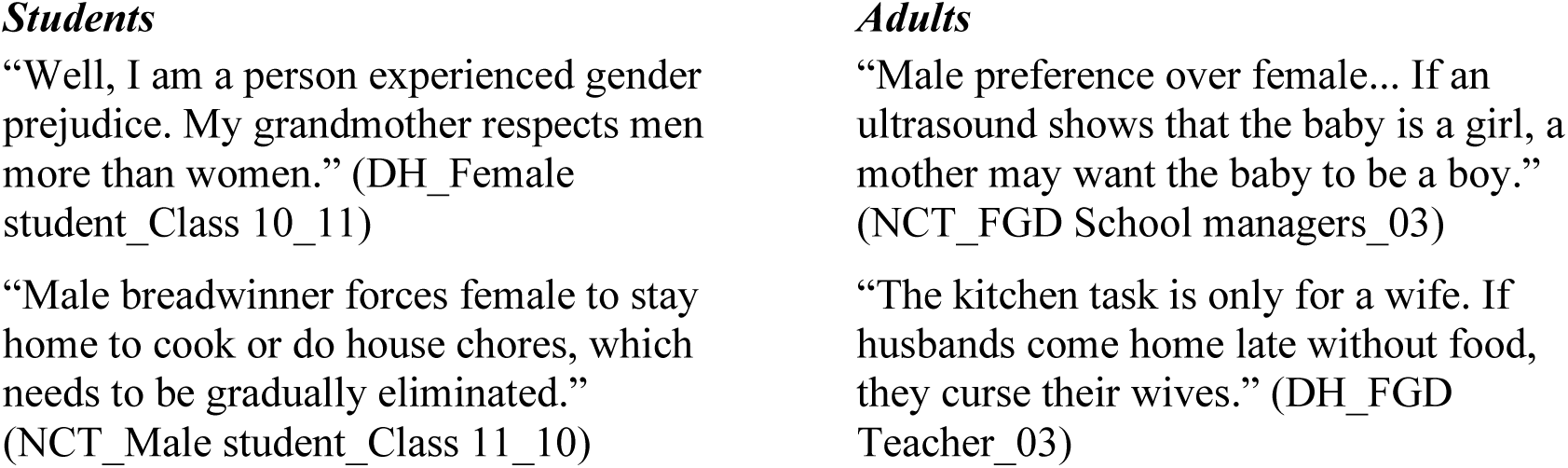

A “bad family environment” was also commonly perceived among many student and adult participants to contribute to violent behavior among students. According to both groups of participants, a “bad family environment may include a lack of family love, children’s exposure to domestic violence (such as husbands who “curse their wives”), direct experience of verbal violence especially by girls or neglect in the family, having divorced parents, and living without parents:

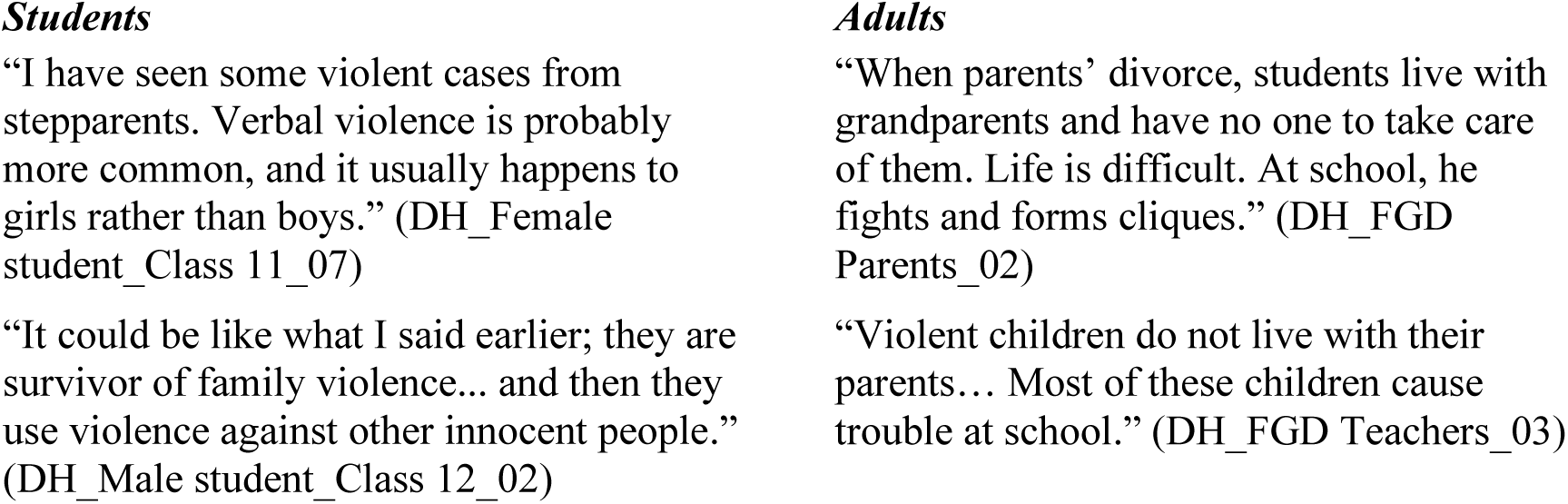

Thus, some student and adult participants perceived that family gender norms— privileging sons and adult male members and relegating female members to subservient roles— and a bad family environment characterized by adversity and violence against children contribute to violence in schools.

**“Outside” high schools** (Figure 1, Box d)

Regarding influences beyond the school environment, some student and adult participants stated that Vietnam’s socio-cultural norms were related to GBV. For example, some participants conveyed that younger generations still are expected to listen to older generations “without argument,” that gender roles should be differentiated, that men and women are seen as unequal, and that men are “superior.” There was a lot of gender prejudice due to the “third gender” (e.g., gay, lesbian, LGBT). These socio-cultural gender norms were seen to contribute to gender discrimination, bullying, and violence:

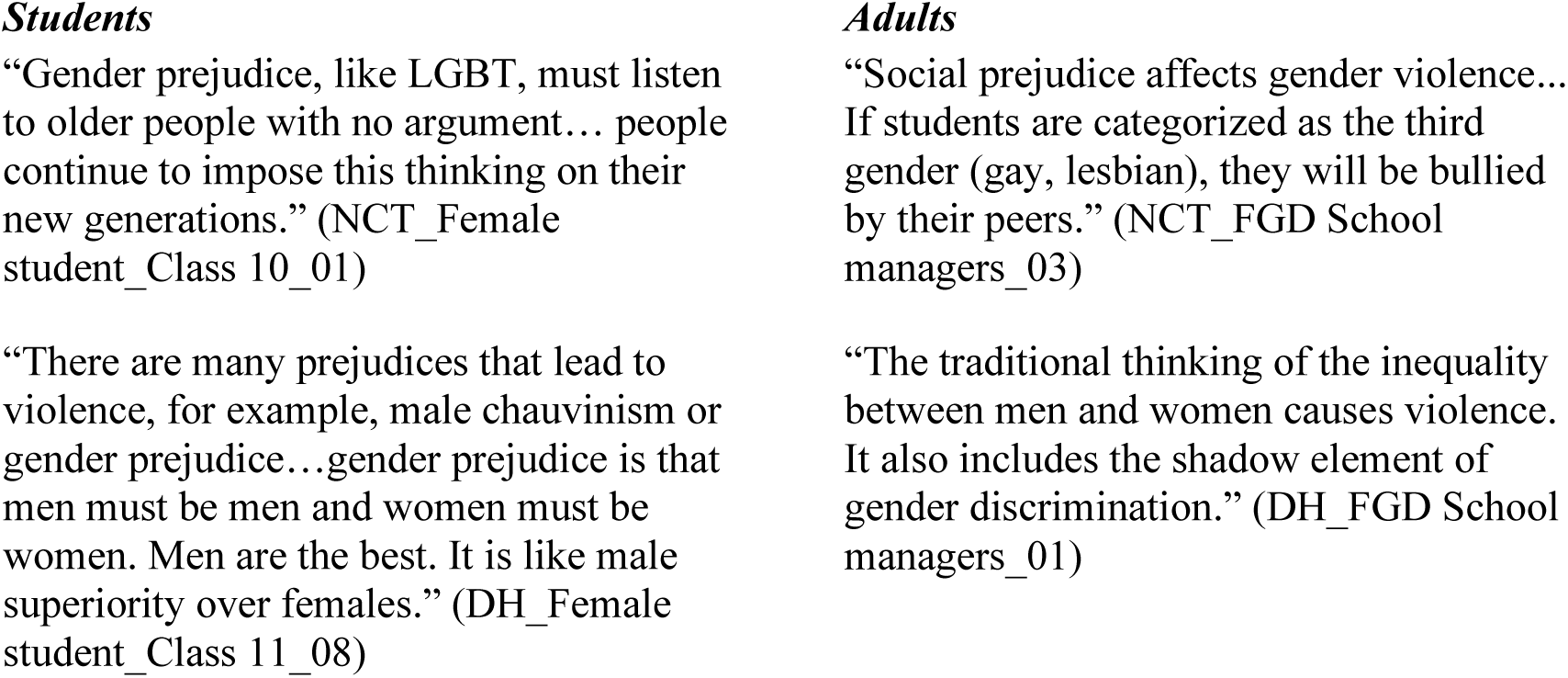

Thus, a common view among student and adult participants was that inequitable socio-cultural gender norms, including specific prejudices against “the third gender,” contributed to gender-based discrimination and violence generally, including in schools.

**Feasible GBV prevention programs and the roles of individuals and society in these programs** (Figure 1, Boxes b to d)

***Feasible future GBV prevention programs***

Some participants suggested maintaining available violence prevention activities at schools for future GBV prevention programs, such as life skills education, role-playing, talk shows on Monday, and short games embedded in the regular school curriculum. Participants also suggested new activities to prevent violence, such as fun games, performing arts, music events, film contests, social media, or using celebrities to promote anti-violence messages in schools.

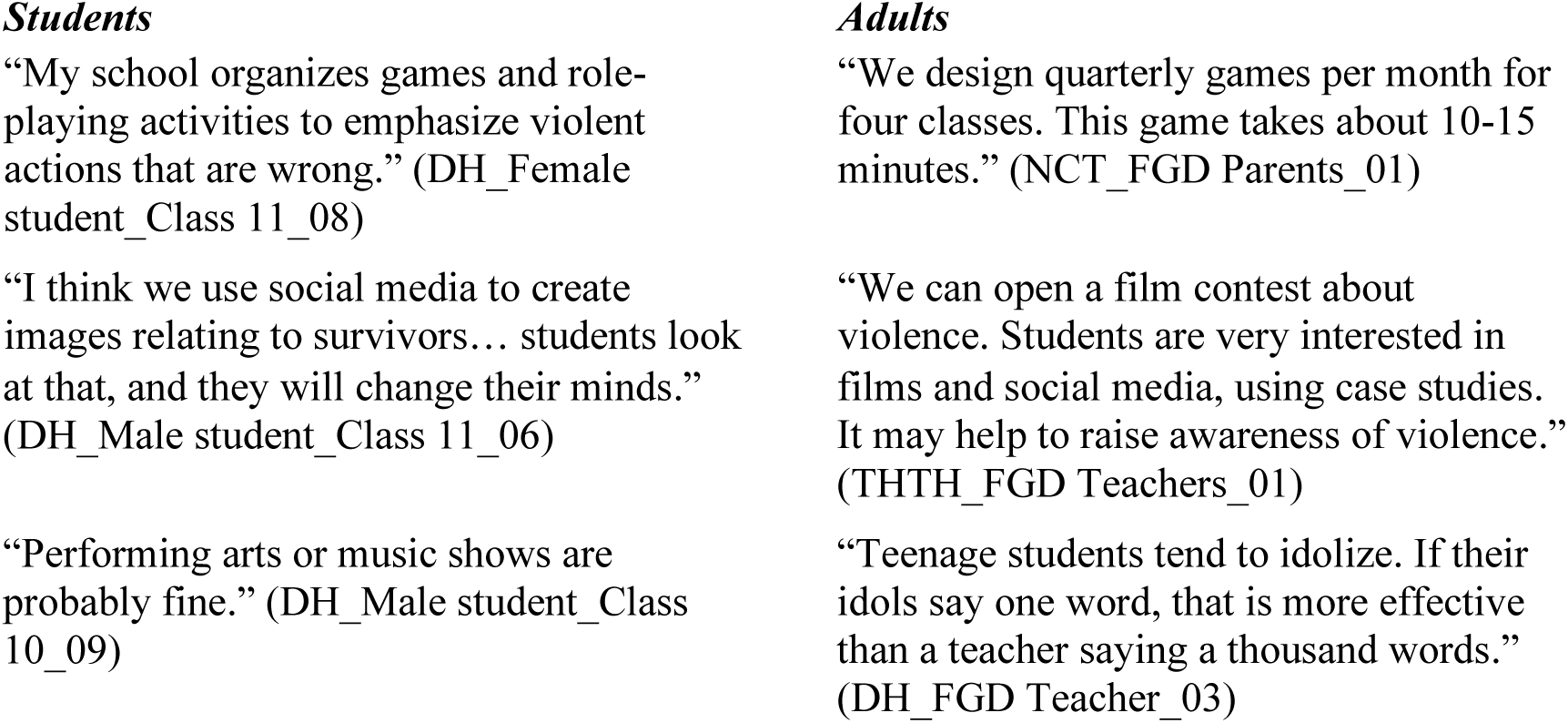

Participants suggested various times to carry out GBV prevention activities, such as 10 to 30 minutes per day, once a month, or every night. To maintain the sustainability of GBV prevention, educational activities should continue in various long-term and engaging forms.

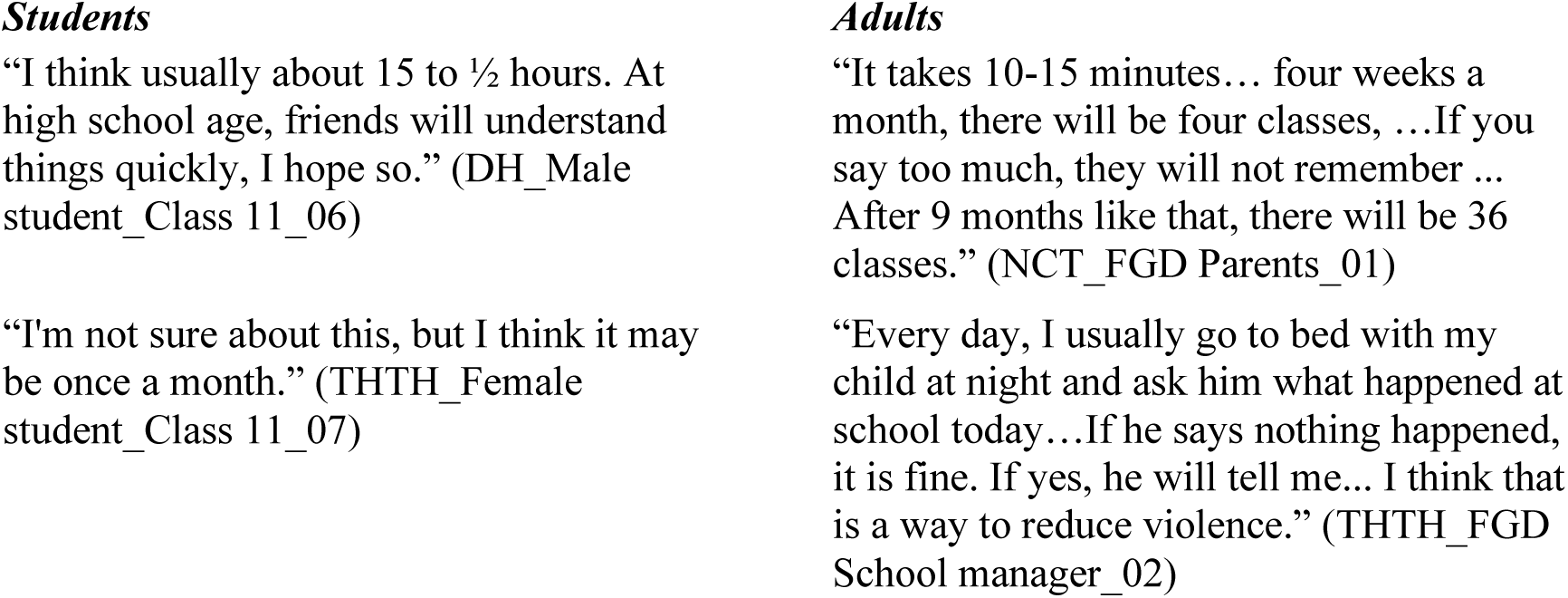

***Roles of communities, schools, and families in GBV prevention***

Most students and adult participants reported that the community, school, and family should work together to prevent and respond to GBV in high schools. Community support could include the enforcement of laws to deter and to prosecute violent offenders in schools. At the school level, participants recommended punishments for violent behavior, installing mailboxes for emergency issues, or posting hotline numbers at schools to encourage reporting cases of GBV and recourse-seeking among GBV survivors.

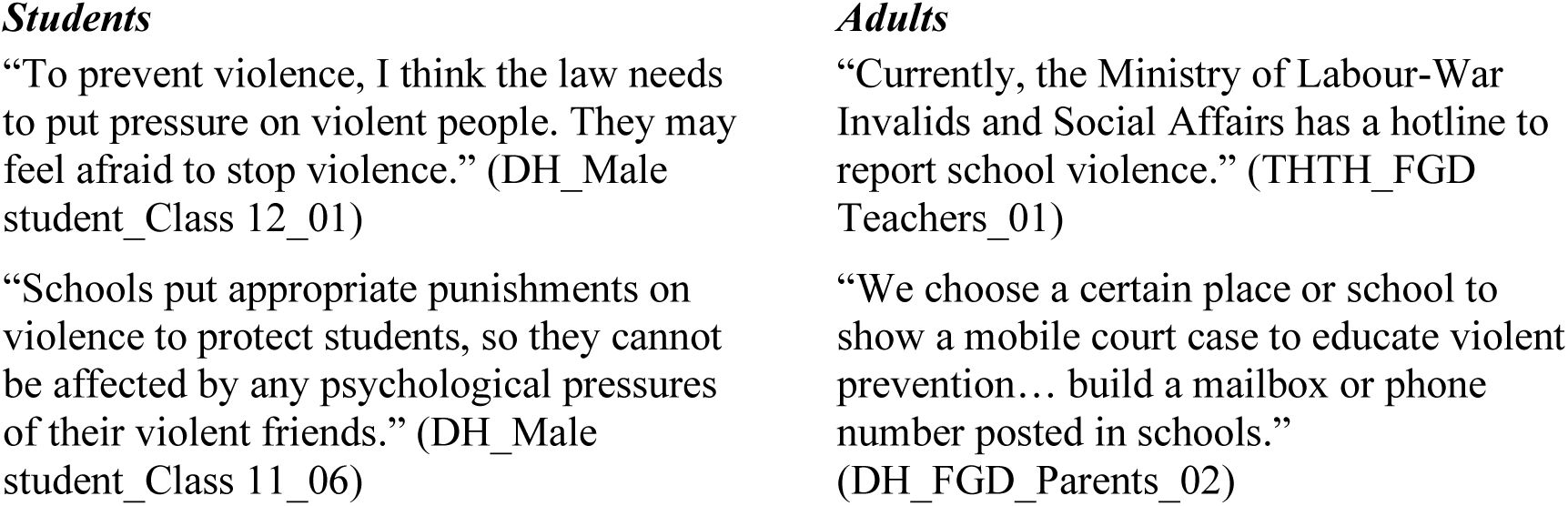

According to many students and adults, the family plays an important role in healthy child development. They suggested that supporting and educating families might help to prevent GBV in high schools. Specifically, parents should “be an example for their children” by avoiding the use of violence with their children, making an effort to understand and to communicate with them about healthy ways to resolve conflicts, and observing them to detect and address violent behaviour:

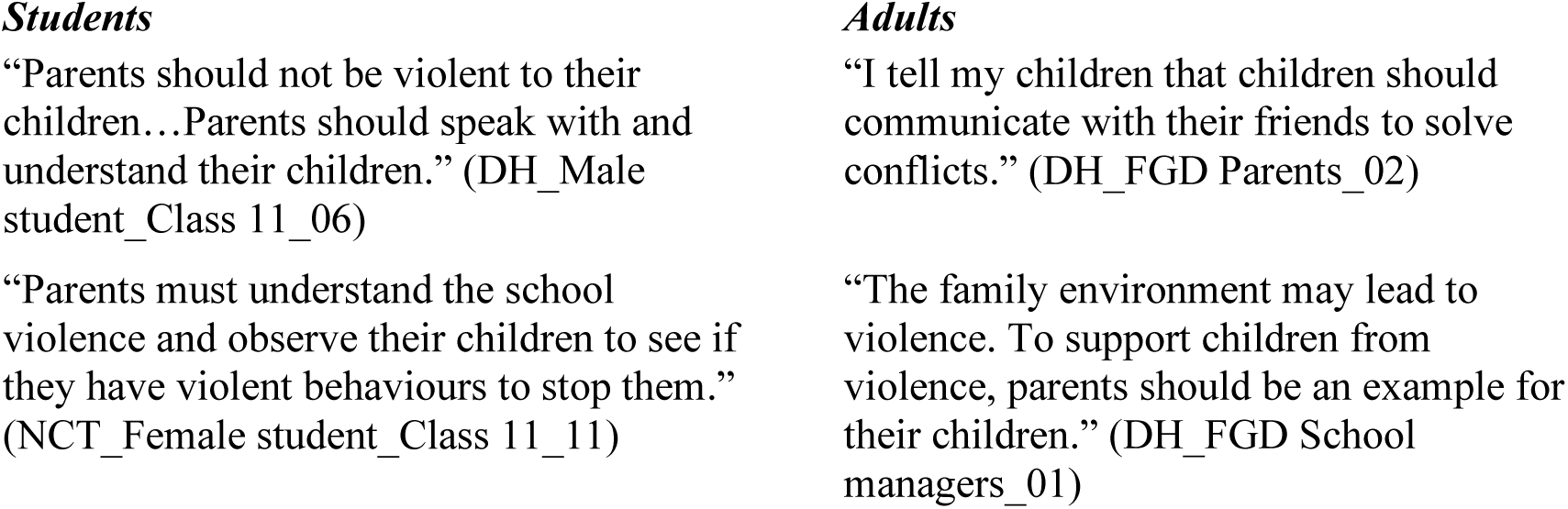

## Discussion

Our study aimed to explore perceptions of GBV as well as acceptable and feasible violence prevention programs at high schools in Vietnam, which are limited (Nguyen et al., 2025). Perceived acts of GBV included physical, psychological, and sexual forms. “Bad blood or impulsive personality” were perceived as perpetrator characteristics. Violent environments, students’ violent behaviour, and cultural norms were the main factors relating to GBV. Future GBV prevention programs for high school students may include famous celebrities, music shows, role-playing, films, or funny games. The community, school, and family should work together to stop violence in high schools.

Findings underscore the multifaceted nature of GBV as perceived by students in high schools, with both physical and psychological violence identified as prevalent forms. Participants widely acknowledged that “violence is an action,” where harm is intentionally enacted by one individual or group against another. Physical violence was characterized by direct aggressive acts such as attacking, punching, kicking, hitting, or pulling hair, which aligns with prior literature describing GBV as involving overt physical harm (Modise & Modise., 2023). Psychological violence emerged as equally salient, encompassing behaviors such as verbal abuse, spreading malicious rumors, and threatening messages on social media. A key insight from the findings is the recognition of social media’s role in amplifying psychological violence, which has emerged as salient in the survey of sexual misconduct in the same schools (Yount et al., 2025) and in prior qualitative interviews with high-school teachers in Vietnam (Anderson et al., 2024). The salient point of psychological violence in this study corroborates prior studies identifying psychological violence as a critical yet often overlooked component of GBV in schools (Akeusola, 2023; Debora & Hasan, 2024).

Moreover, the convergence of views between students and adults suggests a shared understanding of GBV’s complexity, which provides a promising foundation for collaborative intervention strategies. Recognizing both physical and psychological violence, including their digital manifestations, underscores the need for comprehensive, multi-dimensional GBV prevention programs that address school, community, and societal factors (UNICEF, 2020). These findings highlight an urgent need to integrate awareness of GBV’s multifaceted forms into school curricula and involve stakeholders—students, teachers, and parents—in creating supportive environments. Addressing GBV in the digital age requires a dual focus on traditional forms of violence and its emerging dimensions online, an approach supported by international frameworks for combating school violence (UNGEI, 2019).

Physical, psychological, and sexual violence were seen as the most common GBV forms among high school students, which were similar to previous studies (Dogiso et al., 2019; Mingude & Dejene, 2021; Tantu et al., 2020). These GBV forms could lead to students’ self-blame, anxiety, poor school performance, absenteeism, or withdrawal from school (Beyene et al., 2021).

Both students and adults expressed some endorsement of gendered myths about male perpetrators of GBV—that male perpetrators have a *natural* tendency to be violent (Bergenfeld, Lanzas, et al., 2022). Both groups also endorsed certain gendered myths of female perpetrators of GBV—that violent girls were “noisy,” “petty,” or turned small issues into “serious problems.” Other participants focused on specific (and possibly modifiable) characteristics of students who behaved violently, such as impulsivity or the learned tendency to abuse their power with peers like a “tiger mother or father”. Finally, students and adults tended to endorse certain myths that survivors of violence were somehow to blame (Bergenfeld, Lanzas, et al., 2022).—because they were socially “isolated,” unhelpful of their friends, or “too smart,” “too bad,” or “stupid.” Thus, participants endorsed gendered myths about the profiles of those who enacted or experienced violence while commonly asserting that violent behaviour was rooted in the learned desire to control or to exert power over others (Pusch, 2024).

Our study also found that a “bad family environment” (a lack of family love, exposure to domestic violence, being a violent survivor in the family, having divorced parents, and living in a poor family) was perceived to contribute to students’ violent behaviour. These results align with the large body of literature showing that children’s exposure to domestic violence normalizes violent behaviours and fosters emotional distress (Bender et al., 2022), while children’s exposure to adversity more generally also increases psychological stress that can lead to aggression (Xerxa et al., 2020). Additionally, economic hardship often amplifies familial tensions and reduces access to supportive resources, further heightening the risk of youth violence (Peterman et al., 2020). Addressing these factors through schools following the World Health Organization guidelines (World Health Organization, 2022) on parenting interventions to prevent maltreatment and to enhance parent–child relationships may be warranted.

Additionally, our research found that cultural norms (e.g., social, family, or school norms) also related to GBV (Glass et al., 2019; Perrin et al., 2019). These norms reflect the males’ dominant roles, which may be rooted in longstanding notions of male superiority and female inferiority in Vietnam (Thuy Nguyen Thi Thu & Thanh., 2022). According to these longstanding norms, women are expected to complete housework and to care for children, husbands, and elderly people (Samtleben & Müller, 2022); whereas, men are identified as the head of household and “pillar of the house” (An et al., 2022). Female roles are changing in Vietnamese society, but these norms may persist in traditional families and in rural areas of Vietnam (An et al., 2022). Therefore, it is important to raise awareness of the Vietnamese woman’s wider role in society as a means to address gender-based discrimination and harassment (Nhan Dan Journal, 2020), including in schools.

Participants suggested many activities, including games, films, role-playing, or music shows, for future GBV prevention programs. These activities have been identified as promising approaches in prior studies (Bowman et al., 2020; Maya Puspa & Ayu, 2020; Nguyen et al., 2025; Nöcker-Ribaupierre & Wölfl, 2010). Participants also suggested the use of famous celebrities for messaging about GBV prevention, but to our knowledge, no prior studies assessed the effectiveness of using celebrities in violence prevention in high school settings. Despite the high risk of GBV among adolescents in high schools in Vietnam (Tran et al., 2025), no rigorous GBV intervention studies have been undertaken in Vietnamese high schools.

The community, school, and family should participate in any activities to stop violence among high school students. Education is important to increase knowledge and skills for violence prevention (UNESCO, 2024). GBV education can be performed via social media, films, video games, training, counselling, or curriculum-based programs at schools (UNESCO, 2020). The government, local authorities, and school leaders should issue policies or regulations not only to protect students from GBV but also to punish perpetrators (World Health Organization, 2019). Parents take time to teach their children how to prevent violence at school and protect themselves from violence (World Health Organization, 2019).

Few studies have explored perceptions about GBV and acceptable and feasible prevention programs in Vietnam (Yount et al., 2023). The results from the present study offer an in-depth and triangulated understanding of the views of students, parents, teachers, and school managers in high schools. Therefore, these results provide evidence for policymakers to consider appropriate methods for preventing GBV in schools. Moreover, the government can develop a suitable future for GBV intervention to satisfy students’ needs and the expectations of adult constituents. Researchers should pilot culturally suitable programs to ensure their feasibility, acceptability, and safety in the Vietnamese context.

The study has some limitations. The schools were chosen based on rankings and participation in a prior survey of sexual misconduct. As a result, the narratives in this qualitative study may be framed from prior quantitative research in these schools. Schools with different socioeconomic contexts or geographic locations were not included, potentially leaving out crucial variations in GBV perceptions and experiences. While 36 students and 64 adult constituents were included, the sample size may not fully represent the diversity of perspectives across all high schools in the sample, and findings cannot be generalized to all schools in Ho Chi Minh City or Vietnam. The study also excluded participants who did not fully complete the interview. Non-completion may have arisen for a range of reasons that resulted in a loss of important perspectives and saturation of ideas. Participants who completed their interviews and group discussions may have been more inclined to share their thoughts due to their willingness to engage, which may not have reflected the broader population’s views. Although the study aimed to provide culturally relevant insights, strategies for prevention as expressed by participants here may warrant piloting, refinement, and formal testing to ensure they are effective and acceptable across different cultural subgroups.

## Conclusion

Perceptions of GBV and acceptable and feasible violence prevention programs at high schools were explored. Physical, psychological, and sexual violence were noted as salient forms of GBV in high schools. Participants adhered to myths about the “profiles” of students who perpetrate and experience violence while also recognizing the salience of power and control in peer relationships. The primary perceived factors leading to GBV included violent family environments and salient socio-cultural gender norms in families, schools, and broader society. Recommended approaches for GBV intervention among participants included role-plays, films, games, music shows, or the use of celebrities for prevention messaging. Violence prevention among high-school students should involve coordinated efforts in communities, schools, and families. GBV intervention studies are lacking in Vietnamese high schools, so culturally suitable programs should be piloted and formally tested to provide evidence on what works to prevent violence in Vietnamese high schools.

## Data Availability

All data produced in the present work are contained in the manuscript

## Acknowledgments

We thank you, Pham Thi Dan Linh and Vo Y Lam, for helping with the data collection and transcribing. We appreciate Nguyen Nhat Chi Mai for the data transcription.

## Declaration of interest statement

### Funding detail

Emory University of the USA supported this work under Grant [D43TW012188] from the Fogarty International Center of the National Institutes of Health. The primary funder had no role in the research or the decision to publish the findings.

### Authors’ contributions

Discuss proposal and manuscript structure: All authors. Design the interview and focus group discussion guides and contact the three high schools: Hoa H Nguyen. Revise the interview and discussion guides: All authors. Data collection and analysis: Hoa H Nguyen. Revise manuscript: All authors. Approve final version: All authors.

## Short biographical notes

***Hoa Hong Nguyen:*** She was a lecturer at the UMP from 2004 to March 2025. With different roles in health studies, she was one of two principal supervisors of the World Health Organization Survey in 2006. She got special awards, including the Australian Government Scholarship for a master’s degree from Flinders University (2010-2011) and the PhD scholarship from the University of Tasmania, Australia (2015-2020). She is currently working as a university lecturer at Thu Dau Mot University, Binh Duong, Vietnam. Her achievements led to her leadership role as the Vice Head of the Community Health Practice Center (2013) and Head of the Community Health Department (2014-2017). She led the Department of Education Quality Assurance to get the 306th successful AUN-QA assessment of the Public Health Undergraduate Program in 2022. She participated in the ARWIL project (2021-2022) as a leader of the qualitative research team in the STAR-OM project (2020-2023). She received the Australia Award Fellowship, sponsored by DFAT (2023), the VISA D43 Fellowship from the University of North Carolina, USA (2022-2024), and was a member of the International Society for Research on Aggression (2023-2024). She is a Post-doc Fellow of Emory University, USA (2023 -2025).

***Kien G. To:*** Dr. Tô Gia Kiên is a public health researcher and educator. He earned his Bachelor’s degree in Public Health in 2003 from the University of Medicine and Pharmacy at Ho Chi Minh City. He later obtained a Master’s in Public Health from Queensland University of Technology, Australia, in 2009 and a PhD in Public Health from Curtin University, Australia, in 2015. He was promoted to Associate Professor in 2020. Dr. Kiên is the Head of the Department of Health Management and Vice Dean of the Faculty of Public Health at his university, where he has taught since 2004. His research focuses on hospital management, infection control, epidemiology, and healthcare quality. He has worked on many studies related to healthcare accessibility and hospital infection prevention. His expertise includes training healthcare professionals, advising hospitals, and shaping health policies, helping improve healthcare systems through research, education, and leadership.

***Van TH. Hoang:*** Dr. Hoang Thi Hai Van serves as the Head of the Global Health Department at the School of Preventive Medicine and Public Health, Hanoi Medical University. She was previously employed as the deputy head of the Medical Informatics Statistics Department and the Epidemiology Department. In 2000, she obtained a medical doctor degree from Hanoi Medical University. She subsequently pursued a Master of Public Health degree from the University of New South Wales in 2011, a PhD in Public Health from Hanoi Medical University in 2016, and advanced training in Epidemiology and HIV/AIDS research at UCLA in 2017. For more than two decades, Dr. Hoang Thi Hai Van has instructed students at Hanoi Medical University in the fields of biostatistics, research methods, implementation science, epidemiology, global health, and One Health. Additionally, she serves as a research consultant and teaches short courses for various universities, institutions, NGOs, and government agencies in Vietnam. Dr. Van has either led or contributed to several research initiatives in preventive medicine and public health, such as infectious diseases, non-communicable diseases, HIV/AIDS, opioid disorders, violence, etc. She has supervised more than 30 postgraduate students and PhD candidates, and she has authored over 90 scientific publications.

***Lu Gram:*** “Dr Lu Gram is a global health researcher on women’s and communities’ empowerment in low- and middle-income countries and a specialist in collective action for health and gender equality. He is a Sir Henry Wellcome Postdoctoral Fellow at University College London and has published extensively on topics of maternal and child health, women’s empowerment, and community mobilisation.”

***Kathryn M. Yount:*** Dr. Kathryn Yount is Asa Griggs Candler Professor of Global Health (2012) and Professor of Global Health and Sociology (2015) at Emory University. Her research centers on the social determinants of women’s health, including mixed-methods evaluations of social-norms and empowerment-based programs to reduce gender-based violence and health disparities in underserved communities. She has been funded continuously since 2002 from U.S. federal agencies, private foundations, and foreign agencies to work in parts of Asia, Latin America, the Middle East, Sub-Saharan Africa, and Atlanta. These partnerships have contributed 300 <u>publications</u> in the social sciences and global health. Dr. Yount serves as contact PI of the Fogarty/NIH D43 grant <u>CONVERGE</u>, which is training the next generation of science leaders in the prevention of gender-based violence and violence against children in Vietnam and is PI of <u>SCALE</u>, a national implementation trial of an efficacious sexual violence prevention program being implemented with universities across Vietnam. Dr. Yount was a member of the National Academies of Sciences, Engineering, and Medicine, Division of Behavioural and Social Sciences and Education, Committee on Population’s <u>Ad Hoc</u> <u>Committee</u> conducting a Panel Study of Women’s Empowerment, Population Dynamics, and Socioeconomic Development (2022-2024) and is an elected <u>Fellow of the American Academy</u> <u>for the Advancement of Science</u> (AAAS; 2023).

## List of abbreviations

FGDs: Focus group discussions
GBV: Gender-based violence
IDIs: In-depth interviews
PI: Principal investigator

## References

1. Akeusola, B. N. (2023). Social media and the incidence of cyberbullying in Nigeria: Implications for creating a safer online environment. NIU J Humanit, 8(3), 125–137. 10.22373/jai.v9i1.3278

2. An, T. L., Waling, A., & Bourne, A. (2022). Men and masculinities studies in Vietnam: A brief review. Sociol Compass, 16(3), e12965. 10.1111/soc4.12965

3. Anderson, K. M., Macler, A., Bergenfeld, I., Trang, Q. T., & Yount, K. M. (2024). The media and sexual violence among adolescents: Findings from a qualitative study of educators across Vietnam. Arch Sex Behav, 53(6), 2319–2335. 10.1007/s10508-024-02869-7

4. Badriah, S., Tambuala, F., Herlinah, L., Mariani, D., Nurcahyani, L., & Setiawan, H. (2023). The effect of comprehensive sexual education on improving knowledge, attitudes, and skills in preventing premarital sexual behavior in adolescents. Kontakt, 25(1), 404–410. 10.32725/kont.2023.004

5. Bender, A. E., McKinney, S. J., Schmidt-Sane, M. M., Cage, J., Holmes, M. R., Berg, K. A., Salley, J., Bodell, M., Miller, E. K., & Voith, L. A. (2022). Childhood exposure to intimate partner violence and effects on social-emotional competence: A systematic review. J Fam Violence, 37(8), 1263–1281. 10.1007/s10896-021-00315-z

6. Bergenfeld, I., Lanzas, G., Trang, Q. T., Sales, J., & Yount, K. M. (2022). Rape myths among university men and women in vietnam: A qualitative study. J Interpers Violence, 37(3-4), NP1401-NP1431. 10.1177/0886260520928644

7. Bergenfeld, I., Tamler, I., Sales, J. M., Trang, Q. T., Minh, T. H., & Yount, K. M. (2022). Navigating changing norms around sex in dating relationships: A qualitative study of young people in Vietnam. Sex Cult, 26(2), 514–530. DOI:10.1007/s12119-021-09905-x

8. Beyene, A. S., Chojenta, C., Roba, H. S., Melka, A. S., & Loxton, D. (2019). Gender-based violence among female youths in educational institutions of Sub-Saharan Africa: A systematic review and meta-analysis. Syst Rev, 8(1), 59–73. 10.1186/s13643-019-0969-9

9. Beyene, A. S., Chojenta, C. L., & Loxton, D. J. (2021). Consequences of gender-based violence on female high school students in eastern Ethiopia. Afr J Reprod Health, 25(4), 22–33. 10.29063/ajrh2021/v25i4.3

10. Bowman, N. D., Ahn, S. J., & Mercer Kollar, L. M. (2020). The paradox of interactive media: The potential for video games and virtual reality as tools for violence prevention. Frontiers in communication, 5, 580965. 10.3389/fcomm.2020.580965

11. Dan Tri Newspaper. (2024). *Ranking of 10th grade benchmark scores of 108 high schools in Ho Chi Minh City in* 2024. https://dantri.com.vn/giao-duc/xep-hang-diem-chuan-lop-10-cua-108-truong-thpt-tai-tphcm-nam-2024-20240703154607701.htm

12. Debora, C., & Hasan, N. N. N. (2024). Analysis of social media user responses to verbal violence in the cyber world. KOMUNIKA: Jurnal Dakwah dan Komunikasi, 18(1), 85–96. 10.24090.komunika.v18i1.9441

13. Dogiso, A., Shegaze, M., Alagaw, A., & Wassihun, B. (2019). Prevalence and associated factors of gender-based violence among high school female students in Aleta Wondo Town, Southeast Ethiopia. Ethiop J Reprod Health, 11(2), 10. 10.69614/ejrh.v11i2.269

14. Fonteyn, M. E., Vettese, M., Lancaster, D. R., & Bauer-Wu, S. (2008). Developing a codebook to guide content analysis of expressive writing transcripts. Appl Nurs Res, 21(3), 165–168. 10.1016/j.apnr.2006.08.005

15. Glass, N., Perrin, N., Marsh, M., Clough, A., Desgroppes, A., Kaburu, F., Ross, B., & Read-Hamilton, S. (2019). Effectiveness of the Communities Care programme on change in social norms associated with gender-based violence (GBV) with residents in intervention compared with control districts in Mogadishu, Somalia. BMJ Open, 9(3), e023819. 10.1136/bmjopen-2018-023819

16. Grose, R. G., Chen, J. S., Roof, K. A., Rachel, S., & Yount, K. M. (2021). Sexual and reproductive health outcomes of violence against women and girls in lower-income countries: A review of reviews. J Sex Res, 58(1), 1–20. 10.1080/00224499.2019.1707466

17. Grose, R. G., Roof, K. A., Semenza, D. C., Leroux, X., & Yount, K. M. (2019). Mental health, empowerment, and violence against young women in lower-income countries: A review of reviews. Aggress Violent Behav, 46, 25–36. 10.1016/j.avb.2019.01.007

18. Hillis, S., Mercy, J., Amobi, A., & Kress, H. (2016). Global prevalence of past-year violence against children: a systematic review and minimum estimates. Pediatrics, 137(3), e20154079. 10.1542/peds.2015-4079

19. Kathryn M Yount, Yuk Fai Cheong, Irina Bergenfeld, Quach Thu Trang, Jessica M Sales, Yiman Li, & Minh, T. H. (2023). Impacts of GlobalConsent, a web-based social norms edutainment program, on sexually violent behavior and bystander behavior among university men in vietnam: Randomized controlled trial. JMIR Public Health Surveill, 9, e35116. 10.2196/35116

20. Kiger, M. A.-O., & Varpio, L. A.-O. (2020). Thematic analysis of qualitative data: AMEE Guide No. 131. Med Teach, 42(8), 846–854. 10.1080/0142159X.2020.1755030

21. Lange, E., & Young, S. (2019). Gender-based violence as difficult knowledge: Pedagogies for rebalancing the masculine and the feminine. Int J Lifelong Educ, 38(3), 301–326. 10.1080/02601370.2019.1597932

22. Lewis, P., Bergenfeld, I., Thu Trang, Q., Minh, T. H., Sales, J. M., & Yount, K. M. (2022). Gender norms and sexual consent in dating relationships: a qualitative study of university students in Vietnam. Cult Health Sex, 24(3), 358–373. 10.1080/13691058.2020.1846078

23. Lu Gram, Irina Bergenfeld, Katherine M. Anderson, Tran Hung Minh, & Yount, K. (2023). Proximate psychological influences on bystander action to address sexual violence among university men in Vietnam. *PsyArXiv preprints*. 10.31234/osf.io/8ujza

24. Maya Puspa, R., & Ayu, I. (2020). Effectiveness of group counseling role playing techniques to reduce student bullying behavior. Atlantis Press. 10.2991/assehr.k.200814.024

25. Mingude, B. A.., & Dejene, T. M. (2021). Prevalence and associated factors of gender-based violence among Baso high school female students, 2020. Reproductive Health, 18(1), 247. 10.1186/s12978-021-01302-9

26. Ministry of Labour - Invalids and Social Affairs. (2019). Second national study on violence against women in Vietnam 2019 released. https://www.molisa.gov.vn/baiviet/222878?tintucID=222878

27. Modise, J. M., & Modise, P. S. (2023). The fundamental objective of gender-based violence is to address the root causes and motivators of violence against women and girls. Int J Innov Sci Res Technol, 8(5), 2586–2597. 10.5281/zenodo.10057951

28. Nagashima-Hayashi, M. A.-O., Durrance-Bagale, A. A.-O., Marzouk, M., Ung, M., Lam, S. A.-O. X., Neo, P., & Howard, N. A.-O. (2022). Gender-based violence in the Asia-pacific region during COVID-19: A hidden pandemic behind closed doors. Int J Environ Res Public Health, 19(4), 2239. 10.3390/ijerph19042239

29. Nguyen, H. H., To, K. G., Hoang, V. T., Gram, L., & Yount, K. M. (2025). The effectiveness of gender-based violence prevention among adolescents aged 10 to 19 years in Southeast Asia: A systematic review. medRxiv, 2025.2002.2017.25322431. 10.1101/2025.02.17.25322431

30. Nhan Dan Journal. (2020). Enhancing Vietnamese women’s role in society. https://en.nhandan.vn/enhancing-vietnamese-womens-role-in-society-post83845.html

31. Nöcker-Ribaupierre, M., & Wölfl, A. (2010). Music to counter violence: A preventative approach for working with adolescents in schools. Nord J Music Ther, 19, 1–12. 10.1080/08098131.2010.489997

32. Nurdin, Y., Neherta, M., & Meri, D. (2018). The effectiveness of the “Neherta” model as primary prevention of sexual abuse against primary school children in West Sumatera Indonesia 2017. Indian J Public Health Res Dev, 9(10), 446–452. 10.5958/0976-5506.2018.01385.2

33. O’Brien, B. C., Harris Ib Fau - Beckman, T. J., Beckman Tj Fau - Reed, D. A., Reed Da Fau - Cook, D. A., & Cook, D. A. (2014). Standards for reporting qualitative research: A synthesis of recommendations. Acad Med, 89(9), 1245–1251. 10.1097/ACM.0000000000000388

34. Perrin, N., Marsh, M., Clough, A., Desgroppes, A., Yope Phanuel, C., Abdi, A., Kaburu, F., Heitmann, S., Yamashina, M., Ross, B., Read-Hamilton, S., Turner, R., Heise, L., & Glass, N. (2019). Social norms and beliefs about gender based violence scale: a measure for use with gender based violence prevention programs in low-resource and humanitarian settings. Confl Health, 13(1), 6. 10.1186/s13031-019-0189-x

35. Peterman, A., Potts, A., O’Donnell, M., Thompson, K., Shah, N., Oertelt-Prigione, S., & Van Gelder, N. (2020). Pandemics and violence against women and children (Vol. 528). Center for Global Development Washington, DC. https://www.un.org/sexualviolenceinconflict/wp-content/uploads/2020/05/press/pandemics-and-violence-against-women-and-children/pandemics-and-vawg-april2.pdf

36. Pusch, N. (2024). A meta-analytic review of social learning theory and teen dating violence perpetration. J Res Crime & Delinq, 61(2), 171–223. 10.1177/00224278221130004

37. QSR International. (2023). NVivo 14 for Windows.

38. Samtleben, C., & Müller, K.-U. (2022). Care and careers: Gender (in)equality in unpaid care, housework and employment. Res Soc Strat Mobil, 77, 100659. 10.1016/j.rssm.2021.100659

39. Santre, S., & Pumpaibool, T. (2022). Effects of blended learning program for cyber sexual harassment prevention among female high school students in Bangkok, Thailand. Int J Environ Res Public Health, 19(13), 8209. 10.3390/ijerph19138209

40. Tantu, T., Wolka, S., Gunta, M., Teshome, M., Mohammed, H., & Duko, B. (2020). Prevalence and determinants of gender-based violence among high school female students in Wolaita Sodo, Ethiopia: An institutionally based cross-sectional study. BMC Public Health, 20(1), 540. 10.1186/s12889-020-08593-w

41. The Government of Vietnam. (2017a). Decree No. 80/2017/ND-CP on a safe, healthy and friendly education environment which prevents and stops school violence. https://thuvienphapluat.vn/van-ban/EN/Giao-duc/Decree-80-2017-ND-CP-safe-friendly-education-environment-which-prevents-and-stops-school-violence/397496/tieng-anh.aspx

42. The Government of Vietnam. (2017b). Directive No.18/CT-TTg on strengthening measures to prevent and combat violence against children and child abuse. https://thuvienphapluat.vn/van-ban/Van-hoa-Xa-hoi/Chi-thi-18-CT-TTg-2017-ve-tang-cuong-giai-phap-phong-chong-bao-luc-xam-hai-tre-em-349321.aspx

43. Thuy Nguyen Thi Thu, & Thanh, T. L. T. (2022). The female position and role in Korea and Vietnam (An initial comparison). Adult Educ Discourses, 23, 319–331. 10.34768/dma.vi23.633

44. Tran, K. T., Leone, R. M., Swartout, K. M., Tran, M. H., Trinh, O., & Yount, K. M. (2025). Sexual misconduct among high school students in Vietnam. Child Protection and Practice, 5, 100150. 10.1016/j.chipro.2025.100150

45. UN Women. (2017). Access to criminal justice by women subjected to violence in Vietnam. https://vietnam.un.org/sites/default/files/2019-08/Research%20Report%200905.pdf

46. UNESCO. (2020). School-related gender-based violence (SRGBV): a human rights violation and a threat to inclusive and equitable quality education for all. https://unesdoc.unesco.org/ark:/48223/pf0000374509

47. UNESCO. (2024). *What you need to know about ending violence in and through education*. https://www.unesco.org/en/articles/what-you-need-know-about-ending-violence-and-through-education

48. UNFPA Vietnam. (2020). Journey for Change: Second national study on violence against women in Viet Nam 2019. https://vietnam.unfpa.org/en/news/journey-change-second-national-study-violence-against-women-viet-nam-2019-released

49. UNGEI. (2019). Ending school-related gender-based violence. https://www.ungei.org/sites/default/files/Ending-school-related-gender-based-violence-a-series-of-thematic-briefs-2018-eng.pdf

50. UNICEF. (2020). Global status report on preventing violence against children 2020. https://srhr.dspace-express.com/server/api/core/bitstreams/139d446f-c3b2-42f2-ada1-011915a2fe37/content

51. UNICEF. (2021). *Viet Nam SDGCW Survey 2020-*2021. https://www.unicef.org/vietnam/media/8646/file/Adolescents%20findings.pdf

52. UNICEF. (2025). Gender-based violence. https://www.unicef.org/protection/gender-based-violence-in-emergencies

53. World Health Organization. (2019). *School-based violence prevention: A practical handbook* (ISBN: 978-92-4-151554-2). https://www.unicef.org/media/58081/file/UNICEF-WHO-UNESCO-handbook-school-based-violence.pdf

54. World Health Organization. (2022). WHO guidelines on parenting interventions to prevent maltreatment and enhance parent–child relationships with children aged 0–17 years. World Health Organization. https://books.google.com.vn/books?hl=en&lr=&id=Wq2tEAAAQBAJ&oi=fnd&pg=PR6&dq=family+counselling,+parenting+workshops,+and+economic+support+is+crucial+to+mitigating+violence+and+promoting+healthier+family+dynamics+&ots=FkM24OWJEY&sig=Z7js9RwLRi8-OR5YWfRj8xE9hQI&redir_esc=y#v=onepage&q&f=false

55. Wulanyani, N. M. S., Setyani, I., Marheni, A., & Pratama, P. Y. S. (2020). How animated videos and the snakes-ladders game can prevent sexual abuse in children. 399, 202–205. 10.2991/assehr.k.200130.114

56. Xerxa, Y., Rescorla, L. A., Serdarevic, F., Van IJzendorn, M. H., Jaddoe, V. W., Verhulst, F. C., Luijk, M. P., & Tiemeier, H. (2020). The complex role of parental separation in the association between family conflict and child problem behavior. J Clin Child Adolesc Psychol, 49(1), 79–93. 10.1080/15374416.2018.1520118

57. Yount, K. M., Anderson, K. M., Trang, Q. T., & Bergenfeld, I. (2023). Preventing sexual violence in Vietnam: Qualitative findings from high school, university, and civil society key informants across regions. BMC Public Health, 23(1), 1114. 10.1186/s12889-023-15973-5

58. Yount, K. M., Bergenfeld, I., Anderson, K. M., Trang, Q. T., Sales, J. M., Cheong, Y. F., & Minh, T. H. (2022). Theoretical mediators of GlobalConsent: An adapted web-based sexual violence prevention program for university men in Vietnam. Soc Sci Med, 313, 115402. 10.1016/j.socscimed.2022.115402

59. Yount, K. M., Macaulay, M., & Tran, K. T. (2025). Sexually explicit and violent media use among high school students in vietnam: Gender-differentiated links with sexual misconduct victimization, perpetration, and health. medRxiv, 2025.2004.2001.25324943. 10.1101/2025.04.01.25324943

